# The use of personal protective equipment during common industrial hog operation work activities and acute lung function changes in a prospective worker cohort, North Carolina, USA

**DOI:** 10.1101/2020.11.03.20205252

**Authors:** Vanessa R. Coffman, Devon J. Hall, Nora Pisanic, Maya Nadimpalli, Meredith McCormack, Marie Diener-West, Meghan F. Davis, Christopher D. Heaney

## Abstract

**Introduction:** As occupational activities related to acute industrial hog operation (IHO) worker lung function are not well defined, we aimed to identify IHO work activities associated with diminished respiratory function and the effectiveness, if any, of personal protective equipment (PPE) on IHOs.

**Methods:** From 2014-2015, 103 IHO workers were enrolled and followed for 16 weeks. At each bi-weekly visit, lung function measurements were collected via spirometry and work activities and PPE use were self-reported via questionnaire. Generalized linear and linear fixed-effects models were fitted to cross-sectional and longitudinal data.

**Results:** At baseline, increasing years worked on an IHO were associated with diminished lung function, but other activities were less consistent in direction and magnitude. In longitudinal models, only reports of working in feeding/finisher barns, showed a consistent association. However, a −0.3 L (95% confidence interval: −0.6, −0.04) difference in FEV_1_ was estimated when workers wore PPE consistently versus those weeks they did not. In *post-hoc* analyses, we found that coveralls and facemasks were worn less consistently when workers experienced worse barn conditions and had more contact with pigs, but coveralls were worn more consistently as cleaning activities increased.

**Conclusions:** Similar to past studies, baseline estimates were likely obscured by healthy worker bias. Also making it challenging to disentangle the effect of work activities on lung function was the discovery that IHO workers used PPE differently according to work task. These data suggest that interventions may be targeted toward improving barn conditions so that workers can consistently utilize IHO-provided PPE.

**KEY MESSAGES:** *What is already known about this subject?:* Working on industrial hog operations may be deleterious to long- and short-term respiratory health due to airborne bacteria, endotoxin, hazardous gases, dust, and dander in barns. In efficacy studies PPE has been shown to be protective, but studies have shown that PPE utilization among hog workers has historically been sub-optimal.

*What are the new findings?:* As barn conditions worsened and contact with pigs increased, workers in this cohort reported wearing coveralls and face masks less often; however, they reported increased PPE use as they conducted more cleaning activities at work. During weeks when workers wore PPE their lung function declined, a possible cause being the improper use of the equipment leading to a false sense of protection or re-exposure to hazardous contaminants.

*How might this impact on policy or clinical practice in the foreseeable future?:* Given COVID-19, the H1N1 “swine flu” pandemic, our knowledge of antimicrobial resistant pathogens, and increasing awareness about how food systems are linked to the spread of emerging infectious diseases, occupational health intervention research and workplace policies may focus on creating barn environments that are more conducive to PPE use which could help protect workers and consequently the community.

## INTRODUCTION

Swine agricultural workers[1] and veterinarians[2] are at higher risk for adverse respiratory health outcomes than the general public.[3-5] Air sampling studies of swine facilities have reported that exposures include ammonia (NH_3_),[6, 7] hydrogen sulfide (H_2_S) and hundreds of VOCs,[8] dust,[7, 9, 10] endotoxin,[7, 9-11] dander, feed, and microbes,[12, 13] with an alarming proportion of collected bacteria resistant to two or more antibiotics.[14]

Industrial hog operation (IHO) studies that have employed spirometry to measure worker lung function are largely decades old, and most recent investigations rely on self-reported health outcomes. Among the latest U.S. research that employs objective measures of lung function (*i*.*e*., spirometry) is a 1995 study by Schwartz *et al*. that found pig workers were exposed to higher concentrations of dust and had greater work shift declines in forced expiratory volume in the first second (FEV_1_) and forced vital capacity (FVC) than a control group of non-swine farmers.[15] Cross-shift changes in spirometry measurements were also examined by the same research group finding that total dust concentrations ≥2.8 mg/m^3^ were predictive of a ≥10% decline in FEV_1_ and that ammonia concentrations of ≥7.5 ppm were predictive of a ≥3% decline in FEV_1_.[16]

Newer studies focus strongly on chronic obstructive pulmonary disease (COPD) and have not updated our understanding of other health outcomes, even as industrialized pig farming practices evolve. Further, prospective cohort studies are lacking in this population. A systematic review by Douglas *et al*. found that only 2 of 16 published studies of IHO workers were from prospective cohorts.[3] This is not surprising, as previous publications have called for additional research to identify workplace factors that account for the high prevalence of respiratory symptoms in IHO worker populations.[17, 18]

Documenting the deleterious effects of IHOs on the respiratory health of workers in the U.S. has historically been a challenge. Workers’ hesitance to participate in scientific studies for fear of termination, limited access to operations, and corporate influence[19] mean studies are often short-term, lack the ability to determine changes to health on a granular level, and are conducted outside the U.S. This lack of updated data hinders priority setting for the occupational health of the ∼33,000 IHO workers in the States,[19] many of whom are from marginalized communities (e.g., low socioeconomic status, lacking health insurance, minorities, and immigrants).

The work presented here is unique in that by employing fixed-effects regression analyses,[20] workers are compared to themselves. This largely removes confounding from fixed characteristics (*e*.*g*., participant age, sex, race/ethnicity, structural differences in barns, perceived dustiness, and at-home characteristics), which may be present in models estimating the difference in lung function between IHO workers and the general public or other agricultural workers.[15, 21] To the best of our knowledge, this is the first time fixed-effects regression has been used to assess the association between spirometry and IHO work activities and PPE use. In addition, prior studies have examined cross-sectional,[9, 10, 17, 22-24] long-term (*i*.*e*., chronic),[11, 15, 25-27] or cross-shift (*i*.*e*., sub-acute)[16, 21, 26] changes in lung function, but not acute effects.

The purpose of this investigation is to examine the effect a variety of modern IHO work activities and personal protective equipment (PPE) have on lung function changes (*i*.*e*., spirometric measurements) on the acute scale.

## MATERIALS AND METHODS

### Study design

At each of nine study visits, a work practices questionnaire (either baseline or bi-weekly) adapted from the Agricultural Health

Study[https://www.aghealth.nih.gov/collaboration/questionnaires.html] and the American Thoracic Society (ATS)[28] was administered by community investigators. Spirometry was completed at each visit via a Piko-1 machine. At baseline, a reference instrument (Koko spirometer) was also used (**Figure 1**). Each was used in accordance with recognized guidelines[29, 30] and manufacturer’s instructions.[http://www.quickmedical.com/downloads/nspire-piko1_manual.pdf and http://respitechservice.com/Files-hold/KoKo%20User%20Manual.pdf]

**Figure 1.**
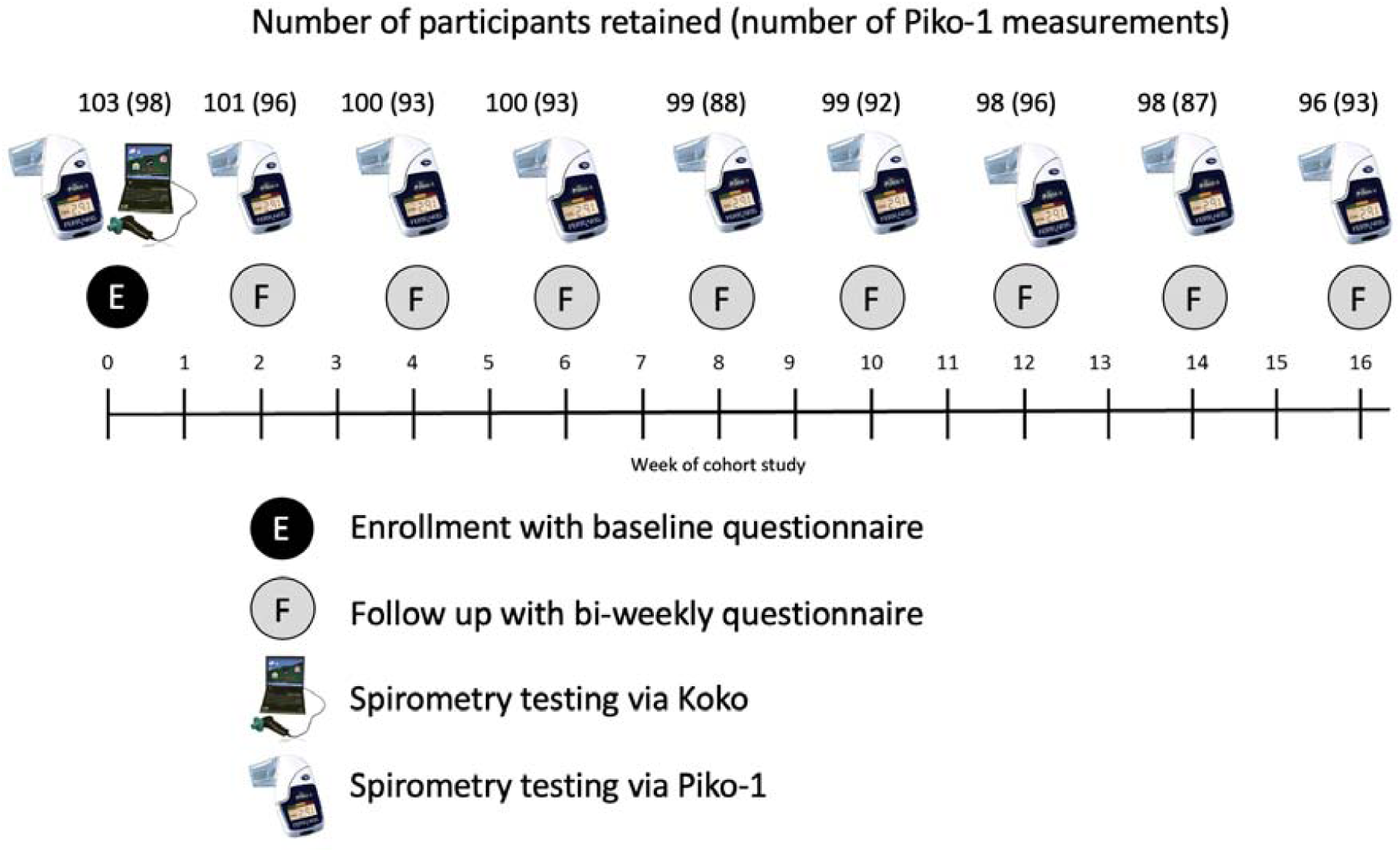
Sampling scheme study visits, participant retention, and measurements obtained within a cohort of industrial hog operation workers, North Carolina, 2013-2014.

### Setting

Participants from hog-producing counties in North Carolina were enrolled as previously detailed.[31] In short, enrollment was conducted on a rolling basis from October 2013 through February 2014. Participants were followed for a maximum of 16 weeks, with a visit from study staff every two weeks. Participants completed a questionnaire and performed spirometry outside of work hours, either at home or in a public place. Data were collected during a Porcine Epidemic Diarrhea Virus (PEDV) epidemic, meaning at times barns were emptied of animals as they either died or were proactively slaughtered.

### Participants

Participants were eligible for inclusion in the sample population if they were current IHO workers (full- or part-time) and agreed to participate in the study. IHO workers were eligible for inclusion in the baseline analysis population if they were enrolled in the study and were eligible for inclusion in the longitudinal analysis population if they completed at least two follow-up visits. Recruitment was performed by Rural Empowerment for Community Help (REACH) personnel using word of mouth and personal connections and powered based on longitudinal analyses. Signed informed consent was obtained from each participant prior to participation. The study protocol was approved by the Johns Hopkins Bloomberg School of Public Health Institutional Review Board.

### Questionnaire

#### Estimates of work practices and conditions

As described in earlier work,[31] all questionnaires were administered by research staff. The enrollment survey instrument was designed to gather data regarding typical work activities and underlying health characteristics. Specific questions were worded as what participants “typically,” “usually,” or “ever” did on-operation and what environmental conditions they were “typically,” “usually,” or “ever” exposed to at work.

At each follow-up visit, information about the frequency, magnitude, and duration of participants’ contact with pigs, job activities, personal behaviors (*e*.*g*., cigarette use), and PPE use was collected. Each question was asked about the week prior. For example, “In the past week have you…”

#### Assessment of lung function

Lung function was assessed by spirometry. A portable Piko-1 pulmonary function device, which records forced expiratory volume in the first second (FEV_1_) and peak expiratory flow rate (PEFr), was used during all study visits by a REACH community data collector. This handheld, portable asthma-tracking tool can be operated without requiring an individual who had completed formal training in National Institute for Occupational Safety and Health (NIOSH)-spirometry to be on-site. Additionally, a Koko machine was employed by a NIOSH-trained coach at baseline. The Koko had a wider range of measurements including: FEV_1_, PEFr, forced vital capacity (FVC), the ratio of forced expiratory volume in the first second to forced vital capacity (FEV_1_/FVC), forced expiratory volume in six seconds (FEV_6_), forced expiratory flow at 25-75% of the pulmonary volume (FEF25-75%), and the ratio of forced expiratory volume in the first second to forced expiratory volume in six seconds (FEV_1_/FEV_6_). Ultimately, in analyses only the Piko-1 measurements and FEV_1_, PEFr, FVC, FEV_1_/FVC Koko measurements were used. Three good trials, as defined by the ATS[ATS] and the NIOSH standards,[30] were attempted for each testing session unless the participant was physically unable to complete the three spirometric maneuvers. The best maneuver per study visit for each person was used in data analysis.

### Data analysis

Baseline data were analyzed via linear regression and linear fixed-effects models were fitted to longitudinal data. Spirometry measurements were modeled as continuous variables, with percent predicted (using reported age, race/ethnicity, and height as predictors in the Hankinson 1999 reference values[32]) at baseline and raw measurements over time.

At baseline, years worked on any IHO and percentage of life working on an IHO were transformed from continuous variables to tertiles due to non-linearity. Average days worked per week and percentage of time at work spent in direct contact with pigs were transformed into binary variables due to non-linearity and skewed distributions. Due to collinearity, reports of workers who ever gave pigs shots and/or antibiotics were combined into a single category.

Drawing on past work in longitudinal analyses,[18] the following unweighted exposure categories and scores were created: (1) *barn conditions* consisting of reports of binary extreme temperature, extreme malodor, extreme dust, vent fans turned off or non-existent at the facility, and a new herd entering the barn(s); (2) *cleaning activities* composed of cleaning chemical use, pesticide use, pressure washing, and torch use; (3) *pig contact*, meaning gave pigs shots and/or gave pigs medicine; and (4) an unweighted sum of all the aforementioned activities with 1 assigned to those reported being done or experienced and 0 to those not reported, for a possible total of 0 to 10. Consistency of PPE use was defined as a worker reporting the gear was worn at least 80% of the time. All exposures were from reports of experiences in the past week. Exposure variables with limited variability (fewer than 10 cases reported over the 752 longitudinal study visits) were *a priori* dropped from analyses to reduce any bias associated with small numbers.

Confounders of interest from the literature and relevant to baseline generalized linear models clustered at the household level included: cigarette smoking, hour of test, and interviewer. Fixed-effects linear regression was used to control for measured and unmeasured confounding in longitudinal analyses. Confounders of interest from the literature and relevant to inclusion in fixed-effects models included: cigarette smoking, hour of test, month of test, and interviewer. A dummy variable was used for hour of test (the time recorded by the Koko machine, when available, or end-of-survey time if not), as this diurnal pattern is not linear.[33]

Spirometry data were analyzed in two ways: (1) all data from the best try (*i*.*e*., no cough, inhale, or delay in start, and a good effort) was used; and (2) as a sensitivity analysis for each model, only measurements with three good tries and two repeatable measurements[29, 30] were used (see **Supplemental Material**). The decision to use all spirometry measurements (regardless of ATS/NIOSH acceptability) as main analyses was based on: (1) Piko-1 devices were not designed to conform to ATS criteria; (2) only the first three maneuvers were recorded using the Piko-1 device (more maneuvers would have met reproducibility had the Koko been used throughout the study); and (3) ATS/NIOSH criteria state that to have a “valid” test both FEV_1_ and FVC measures must have three good tries and both must have two measurements within 0.15 L of each other. Since the Piko-1 device does not assess FVC, this would have made even acceptable FEV_1_ measurements technically invalid.

Crude analyses are reported as main analyses, while fully adjusted models are shown as sensitivity analyses because: (1) fixed-effects models do not suffer from the same necessity for adjustment as other random effects models; and (2) baseline models are unable to converge with many adjustments for confounders.

Data was analyzed using Stata (StataCorp. 2017. *Stata* Statistical Software: Release 15. College Station, TX: *StataCorp* LP).

## RESULTS

At baseline, 103 workers entered the cohort, giving 98 FEV_1_ measurements via Piko-1 and 90 workers remained enrolled in the final week and provided 87 measurements (**Figure 1**). In examining key characteristics stratified by those who were able to complete spirometry tests at baseline and those who were not, we saw little difference except that more non-smokers than smokers were able to provide Koko maneuvers and overall more participants were able to provide Piko-1 measurements (98 of 103) than Koko measurements (69 of 103) (**Table 1**).

**Table 1.** Differences in population characteristics stratified by available baseline spirometry data from two instruments during enrollment into an industrial hog operation worker cohort.

| Characteristic | Koko measurement |  | Piko-1 measurement |  |
| --- | --- | --- | --- | --- |
|  | Has | Does not have | Has | Does not have |
| Workers in cohort, *n* | 69 | 34 | 98 | 4 |
| Age in years, *mean (SD)* | 39 (11) | 35 (9) | 38 (11) | 25 (-) |
| Sex, *n (%)* |  |  |  |  |
| Male | 34 (49) | 21 (66) | 54 (55) | 1 (50) |
| Female | 35 (51) | 11 (34) | 45 (45) | 1 (50) |
| Race/ethnicity, *n (%)* |  |  |  |  |
| Hispanic, non-black | 68 (100) | 20 (63) | 88 (90) | 0 (0) |
| Black | 0 (0) | 12 (34) | 10 (10) | 2 (100) |
| Education status, *n (%)* |  |  |  |  |
| Less than high school education | 34 (51) | 13 (41) | 49 (51) | 0 (0) |
| High school degree/GED or college | 33 (49) | 19 (59) | 47 (49) | 3 (100) |
| Height (cm), *mean (SD)* | 165 (9) | 166 (14) | 165 (11) | - |
| Weight (lbs), *mean (SD)* | 172 (31) | 174 (35) | 172 (32) | - |
| Body mass index, *mean (SD)* | 29 (5) | 29 (6) | 29 (5) | - |
| Used a gym or workout facility in the last three months, *n (%)* |  |  |  |  |
| Yes | 4 (6) | 5 (16) | 8 (8) | 1 (50) |
| No | 65 (94) | 27 (84) | 91 (92) | 1 (50) |
| Current cigarette smoker, *n (%)* |  |  |  |  |
| Yes | 5 (7) | 8 (73) | 12 (16) | 1 (100) |
| No | 62 (93) | 3 (37) | 65 (84) | 0 (0) |
| Had health insurance, *n (%)* |  |  |  |  |
| Yes | 34 (50) | 14 (44) | 46 (47) | 2 (100) |
| No | 34 (50) | 18 (56) | 52 (53) | 0 (0) |
| Does not seek medical care under any circumstance, *n (%)* |  |  |  |  |
| Yes | 4 (6) | 0 (0) | 4 (4) | 0 (0) |
| No | 63 (94) | 32 (100) | 93 (96) | 2 (100) |
| Auto repair or use of chemicals outside of work, *n (%)* |  |  |  |  |
| Yes | 3 (4) | 3 (10) | 6 (6) | 0 (0) |
| No | 64 (96) | 28 (90) | 91 (94) | 1 (100) |
| Had any pet that goes inside the home, *n (%)* |  |  |  |  |
| Yes | 15 (39) | 3 (33) | 18 (38) | - |
| No | 23 (61) | 6 (64) | 29 (62) | - |
| Lived on same property as an IHO, *n (%)* |  |  |  |  |
| Yes | 7 (11) | 1 (3) | 8 (8) | 0 (0) |
| No | 58 (89) | 31 (97) | 87 (92) | 2 (100) |
| Interviewer, *n (%)* |  |  |  |  |
| A | 0 (0) | 2 (8) | 2 (2) | 0 (0) |
| B | 37 (55) | 6 (25) | 41 (47) | 2 (67) |
| C | 28 (42) | 8 (33) | 35 (40) | 1 (33) |
| D | 2 (3) | 8 (33) | 10 (11) | 0 (0) |

Note. SD = standard deviation. IHO = industrial hog operation.

The concordance between best Piko-1 and Koko measurements differed by test (R^2^: 0.79 for FEV_1_ measures and R^2^: 0.40 for PEFr measures) (**Table 2**). Using Bland-Altman plots, Koko FEV_1_ values were greater than Piko-1 for the same individuals; however, Piko-1 values for PEFr were greater than paired Koko values (data not shown). Due to the low agreement for PEFr measures, we chose to only use FEV_1_ measures in longitudinal analyses.

**Table 2.**
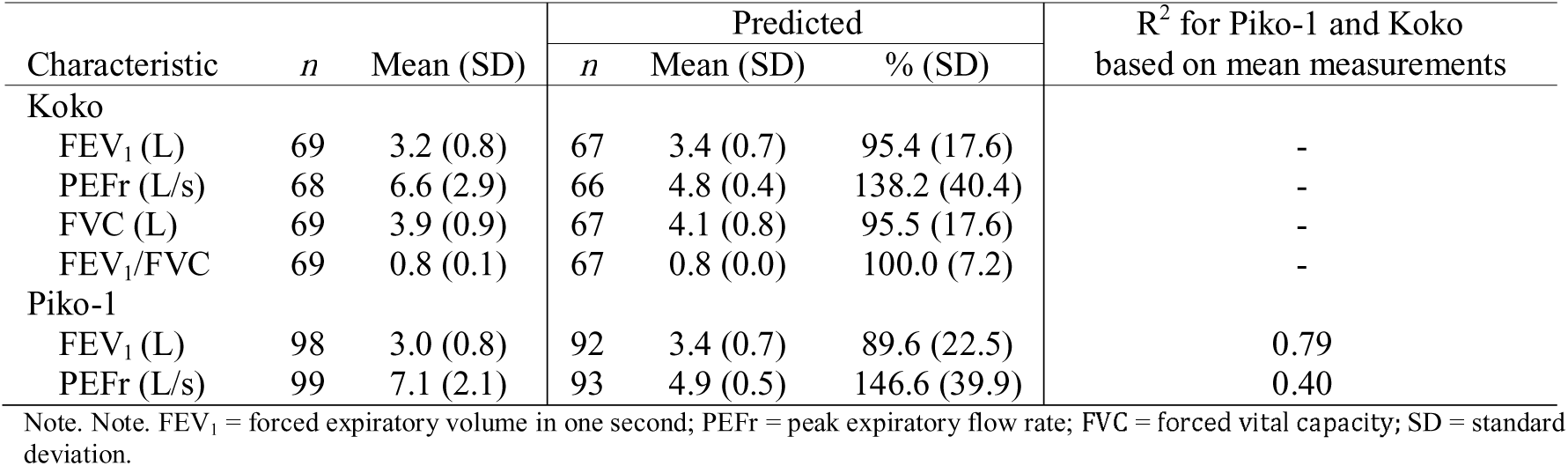
Descriptive statistics of all spirometry measurements at baseline within an industrial hog operation worker cohort, North Carolina, 2013-2014.

| Characteristic | *n* | Mean (SD) | Predicted |  |  | R<sup>2</sup> for Piko-1 and Koko<br>based on mean measurements |
| --- | --- | --- | --- | --- | --- | --- |
|  |  |  | *n* | Mean (SD) | % (SD) |  |
| Koko |  |  |  |  |  |  |
| FEV<sub>1</sub> (L) | 69 | 3.2 (0.8) | 67 | 3.4 (0.7) | 95.4 (17.6) | - |
| PEFr (L/s) | 68 | 6.6 (2.9) | 66 | 4.8 (0.4) | 138.2 (40.4) | - |
| FVC (L) | 69 | 3.9 (0.9) | 67 | 4.1 (0.8) | 95.5 (17.6) | - |
| FEV<sub>1</sub>/FVC | 69 | 0.8 (0.1) | 67 | 0.8 (0.0) | 100.0 (7.2) | - |
| Piko-1 |  |  |  |  |  |  |
| FEV<sub>1</sub> (L) | 98 | 3.0 (0.8) | 92 | 3.4 (0.7) | 89.6 (22.5) | 0.79 |
| PEFr (L/s) | 99 | 7.1 (2.1) | 93 | 4.9 (0.5) | 146.6 (39.9) | 0.40 |

Note. Note. FEV<sub>1</sub> = forced expiratory volume in one second; PEFr = peak expiratory flow rate; FVC = forced vital capacity; SD = standard deviation.

Measured lung function was similar to predicted values (**Table 2**) and very few participants had obstructive or restrictive lung disease (**Table S1**), as classified either by the Global Initiative for Chronic Obstructive Lung Disease (GOLD) criteria[34] or by using the lower limit of normal (LLN) as a cutoff for healthy versus non-healthy lung function.

Relationships between the most reported symptoms at enrollment and percent predicted spirometry values were examined to assess the need to include symptoms as confounders in subsequent models (**Table S2** and **S3**). Only report of doctor-diagnosed asthma (a static characteristic) was significantly associated with lung function decline and so symptoms were not included.

Ten binary self-reported measures of exposure and two variables transformed into tertiles were examined in association with baseline measures of lung function (**Table 3)**.

**Table 3.** Crude baseline relationship between reported work exposures and measured lung function within an industrial hog operation worker cohort, North Carolina, 2013-2014 using GLM and clustered at the household level.

| Characteristic | *n* | % Predicted FEV<sub>1</sub><sup>a</sup> |  | % Predicted FVC<sup>a</sup> |  | % Predicted FEV<sub>1</sub>/FVC<sup>a</sup> |  | *n* | % Predicted FEV<sub>1</sub><sup>b</sup> |  |
| --- | --- | --- | --- | --- | --- | --- | --- | --- | --- | --- |
|  |  | β (95% CI) | *p*-value | β (95% CI) | *p*-value | β (95% CI) | *p*-value |  | β (95% CI) | *p*-value |
| Have you ever |  |  |  |  |  |  |  |  |  |  |
| Given pigs shots and/or antibiotics | 65 | -0.3 (-9.5, 8.9) |  | -3.0 (-11.5, 5.5) |  | 3.4 (-0.2, 7.1) |  | 90 | 5.7 (-4.3, 15.7) |  |
| Drawn pig blood | 65 | 5.0 (-19.8, 29.8) |  | 1.1 (-18.4, 20.6) |  | 2.1 (-2.8, 7.0) |  | 90 | 6.3 (-16.3, 29.0) |  |
| Handled pig manure | 65 | 0.7 (-7.5, 9.0) |  | 0.1 (-8.7, 8.9) |  | 0.4 (-2.9, 3.7) |  | 85 | -1.9 (-11.5, 7.8) |  |
| Applied pesticides in or around the barns | 65 | 0.01 (-8.0, 8.1) |  | -4.3 (-12.7, 4.1) |  | 4.3 (0.9, 7.7) |  | 90 | -1.0 (-10.3, 8.3) |  |
| Washed work clothes with household laundry | 64 | 0.8 (-10.7, 12.2) |  | -1.0 (12.3, 10.3) |  | 1.4 (-2.5, 5.3) |  | 88 | 3.1 (-13.0, 19.2) |  |
| Do you typically |  |  |  |  |  |  |  |  |  |  |
| Work exclusively in sow, nursery, and/or farrow barns | 63 | 2.0 (-6.8, 10.8) |  | 0.5 (-8.3, 9.4) |  | 2.0 (-1.1, 5.0) |  | 87 | 4.4 (-7.1, 16.0) |  |
| Work exclusively in feeder and/or finisher barns | 63 | -3.9 (-12.7, 5.0) |  | -2.4 (-11.3, 6.5) |  | -1.8 (-5.9, 2.3) |  | 80 | -4.6 (-10.3, 1.2) |  |
| Always wear a mask and bodysuit and eye protection | 64 | 4.9 (-4.1, 13.9) |  | 4.5 (-6.5, 15.5) |  | 0.3 (-3.7, 4.3) |  | 89 | 9.1 (-5.1, 23.4) |  |
| Work 7 days per week | 65 | 0.7 (-7.4, 8.9) |  | 2.1 (-6.2, 10.3) |  | -1.7 (-4.7, 1.3) |  | 89 | 0.6 (-8.4, 9.6) |  |
| 100% of time at work spent in direct contact with hogs | 64 | -6.7 (-15.5, 2.2) |  | -7.0 (-15.7, 1.8) |  | 0.5 (-3.2, 4.2) |  | 87 | -9.4 (-19.5, 0.6) |  |
| Years worked on any IHO | 62 |  |  |  |  |  |  | 80 |  |  |
| Tertile 1 (1-5 years) |  | Ref (0.0) |  | Ref (0.0) |  | Ref (0.0) |  |  | Ref (0.0) |  |
| Tertile 2 (6-10 years) |  | -10.3 (-19.9, -0.6) |  | -10.1 (-20.2, -0.002) |  | -0.7 (-5.8, 4.4) |  |  | -6.4 (-18.0, 5.2) |  |
| Tertile 3 (11-27 years) |  | -9.8 (-19.3, -0.2) |  | -10.8 (-20.6, -1.0) |  | 0.6 (-2.6, 3.7) |  |  | -8.9 (-20.1, 2.3) |  |
| Trend |  |  | 0.04 |  | 0.03 |  | 0.7 |  |  | 0.12 |
| Percent of life working on any IHO | 62 |  |  |  |  |  |  | 80 |  |  |
| Tertile 1 (2.4-11.6%) |  | Ref (0.0) |  | Ref (0.0) |  | Ref (0.0) |  |  | Ref (0.0) |  |
| Tertile 2 (11.7-26.3%) |  | -12.4 (-22.8, -1.9) |  | -9.1 (-20.7, 2.5) |  | -4.3 (-8.5, -0.1) |  |  | -14.9 (-27.5, -2.2) |  |
| Tertile 3 (26.4-51.9%) |  | -6.5 (-18.2, 5.3) |  | -5.8 (-17.6, 6.0) |  | -1.2 (-5.2, 2.7) |  |  | -12.2 (-24.3, -0.1) |  |
| Trend |  |  | 0.3 |  | 0.4 |  | 0.6 |  |  | 0.05 |

Note. FEV<sub>1</sub> = forced expiratory volume in one second; FVC = forced vital capacity; CI = Confidence interval. IHO = industrial hog operation.
a. Performed on a Koko spirometer.
b. Performed on a Piko-1 spirometer.

In this crude analysis, 8 of 56 associations were statistically significant (*i*.*e*., confidence interval did not include the null), with one relationship (FEV_1_/FVC and pesticide application) slightly significant but in the unhypothesized direction. Worse lung function was seen in those who worked on any IHO longer than those in the first tertile, with significant associations for trends in 3 of 8 models (**Table 3**). In sensitivity analyses, adjusting for the hour of spirometric test, current cigarette smoking, and interviewer, the relationship between pesticide application and FEV_1_/FVC strengthened (**Table S4**), while the associations between length of time working on any IHO weakened, although 14 of 16 remained in the negative direction.

In longitudinal analyses, limited changes in respiratory function were observed between weeks when an activity was versus was not performed (**Table 4**).

**Table 4.** Relationship between binary exposure activities and spirometry measurements over time within an industrial hog operation worker cohort, North Carolina, 2013-2014.

| Characteristic in the past week | Crude FEV<sub>1</sub> (L) |  | Adjusted FEV<sub>1</sub><sup>a</sup> (L) |  |
| --- | --- | --- | --- | --- |
|  | visits (workers)<sup>b</sup> | β (95% CI) | visits (workers)<sup>b</sup> | β (95% CI) |
| Experienced some form of dustiness or odor<sup>c</sup> | 700 (99) | -0.1 (-0.2, 0.1) | 691 (99) | -0.02 (-0.2, 0.1) |
| Performed a cleaning activity<sup>d</sup> | 720 (99) | 0.1 (-0.1, 0.2) | 711 (99) | 0.01 (-0.1, 0.2) |
| Had pig contact<sup>e</sup> | 718 (100) | -0.1 (-0.2, 0.1) | 708 (100) | -0.1 (-0.3, 0.1) |
| Performed two or three of the above<sup>f</sup> | 687 (99) | 0.02 (-0.1, 0.2) | 679 (99) | 0.03 (-0.1, 0.2) |
| Used any protection consistently<sup>g</sup> | 712 (100) | -0.3 (-0.6, -0.1) | 703 (100) | -0.3 (-0.6, -0.04) |
| Handwashing at least 8 times per shift<sup>h</sup> | 717 (100) | 0.1 (-0.1, 0.2) | 707 (100) | 0.1 (-0.1, 0.2) |
| Worked 7 days | 738 (101) | -0.1 (-0.2, 0.1) | 726 (100) | -0.1 (-0.2, 0.1) |
| Worked at least 45 hours | 728 (101) | 0.004 (-0.1, 0.1) | 717 (100) | -0.1 (-0.2, 0.1) |

Note. FEV<sub>1</sub> = forced expiratory volume in one second; CI = confidence interval. All estimates were made using fixed-effects regression. *a.* Hour of test (dummy), month of test (dummy), smoked in the past 12 hours (binary), and interviewer (dummy)-adjusted relationship between reported symptoms and spirometry measurements over time within an industrial hog operation worker cohort, North Carolina, 2013-2014 using fixed-effects regression. *b.* The number of observations equals the number of individual visits (1-8) for the number of persons (*i.e.*, groups) with both a response to the symptom question and a Piko-1 spirometry test result. *c.* Reported at least one of the following: extreme temperature, extreme malodor, extreme dust, vents off, or a new herd entering the barns *d.* Reported at least one of the following: used cleaning chemicals and/or pesticides, pressure washed or used a torch *e.* Reported at least one of the following: gave hogs shots and/or medicine. *f.* Binary (yes/no) to a, b, and/or c. *g.* Consistently (≥80% of the time at work) wore at least one of the following: mask, glasses, or bodysuit/coveralls. *h.* 8 is the median.

However, PPE use was statistically significant in the un-hypothesized adverse direction in both crude and adjusted models and when exposure was either collapsed into binary measures (**Table 4**) or expanded to detail the number of activities performed (**Table S5**).

To further explore the relationship between PPE use and diminished lung function of this un-hypothesized association, we *post-hoc* chose to examine the association between type of PPE use and lung function using fixed-effects linear regression (**Table S6**). While consistent face protection usage (defined as at least 80% mask and/or eye protection in the past week) showed negligible effects on respiratory function, the consistent use of body protection (defined as at least 80% coverall/full body suit utilization in the past week) was associated with 0.2 to 0.3 L declines in FEV_1_ in unadjusted and adjusted models respectively (crude β: −0.2 L; 95% CI: −0.4, −0.03; adjusted β: −0.3 L; 95% CI: −0.6, −0.1) (**Table S6**).

*Post-hoc* we also explored which workers were more likely to wear certain PPE. Pearson chi-squared tests for associations between on-IHO work activities and PPE use (**Table 5**) and reported symptoms and PPE use were employed (**Table S7**).

**Table 5.**
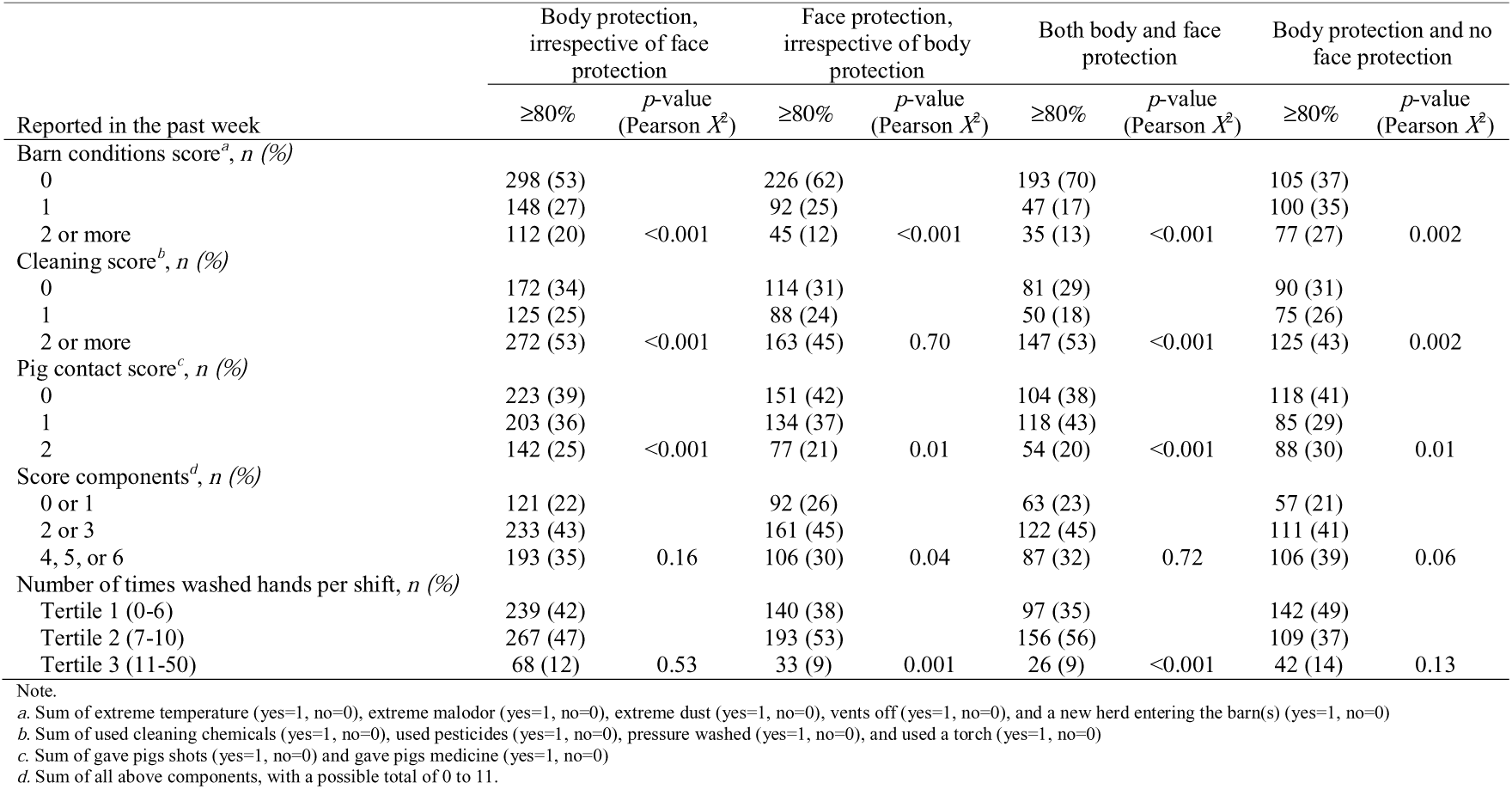
Cross tabulations of personal protective equipment and reported work exposures within an industrial hog operation worker cohort, North Carolina, 2013-2014.

| Reported in the past week | Body protection,<br>irrespective of face<br>protection |  | Face protection,<br>irrespective of body<br>protection |  | Both body and face<br>protection |  | Body protection and no<br>face protection |  |
| --- | --- | --- | --- | --- | --- | --- | --- | --- |
| | ≥80% | *p*-value<br>(Pearson $X^2$) | ≥80% | *p*-value<br>(Pearson $X^2$) | ≥80% | *p*-value<br>(Pearson $X^2$) | ≥80% | *p*-value<br>(Pearson $X^2$) |
| Barn conditions score<sup>a</sup>, *n* (%) |  |  |  |  |  |  |  |  |
| 0 | 298 (53) |  | 226 (62) |  | 193 (70) |  | 105 (37) |  |
| 1 | 148 (27) |  | 92 (25) |  | 47 (17) |  | 100 (35) |  |
| 2 or more | 112 (20) | <0.001 | 45 (12) | <0.001 | 35 (13) | <0.001 | 77 (27) | 0.002 |
| Cleaning score<sup>b</sup>, *n* (%) |  |  |  |  |  |  |  |  |
| 0 | 172 (34) |  | 114 (31) |  | 81 (29) |  | 90 (31) |  |
| 1 | 125 (25) |  | 88 (24) |  | 50 (18) |  | 75 (26) |  |
| 2 or more | 272 (53) | <0.001 | 163 (45) | 0.70 | 147 (53) | <0.001 | 125 (43) | 0.002 |
| Pig contact score<sup>c</sup>, *n* (%) |  |  |  |  |  |  |  |  |
| 0 | 223 (39) |  | 151 (42) |  | 104 (38) |  | 118 (41) |  |
| 1 | 203 (36) |  | 134 (37) |  | 118 (43) |  | 85 (29) |  |
| 2 | 142 (25) | <0.001 | 77 (21) | 0.01 | 54 (20) | <0.001 | 88 (30) | 0.01 |
| Score components<sup>d</sup>, *n* (%) |  |  |  |  |  |  |  |  |
| 0 or 1 | 121 (22) |  | 92 (26) |  | 63 (23) |  | 57 (21) |  |
| 2 or 3 | 233 (43) |  | 161 (45) |  | 122 (45) |  | 111 (41) |  |
| 4, 5, or 6 | 193 (35) | 0.16 | 106 (30) | 0.04 | 87 (32) | 0.72 | 106 (39) | 0.06 |
| Number of times washed hands per shift, *n* (%) |  |  |  |  |  |  |  |  |
| Tertile 1 (0-6) | 239 (42) |  | 140 (38) |  | 97 (35) |  | 142 (49) |  |
| Tertile 2 (7-10) | 267 (47) |  | 193 (53) |  | 156 (56) |  | 109 (37) |  |
| Tertile 3 (11-50) | 68 (12) | 0.53 | 33 (9) | 0.001 | 26 (9) | <0.001 | 42 (14) | 0.13 |

Note.
a. Sum of extreme temperature (yes=1, no=0), extreme malodor (yes=1, no=0), extreme dust (yes=1, no=0), vents off (yes=1, no=0), and a new herd entering the barn(s) (yes=1, no=0)
b. Sum of used cleaning chemicals (yes=1, no=0), used pesticides (yes=1, no=0), pressure washed (yes=1, no=0), and used a torch (yes=1, no=0)
c. Sum of gave pigs shots (yes=1, no=0) and gave pigs medicine (yes=1, no=0)
d. Sum of all above components, with a possible total of 0 to 11.

While symptoms were not meaningfully associated with different levels of PPE utilization, work activities were. Reports of consistent usage of body protection (irrespective of face protection and with non-consistent face protection) as well as the use of both types of PPE increased during weeks when the most cleaning activities were reported (**Table 5**). Notably, body and face protection use (irrespective of the other type of PPE) declined as reports of barn conditions worsened and as contact with pigs increased, as did the use of both types of protection.

## DISCUSSION

While our original analyses yielded little meaningful results regarding IHO workplace activities and within-person changes in lung function, our *post hoc* analyses of PPE usage provide a view of how IHO work conditions relate to PPE utilization. To the best of our knowledge this is the first time an analysis of U.S. IHO work conditions and PPE use by day-to-day workers has been presented; aiding our understanding of why PPE usage by IHO workers may be sub-optimal.[35-37]

### Baseline analyses demonstrate the healthy worker effect

At baseline, reports of time (years and percent of life) worked on any IHO were associated with clinically relevant (∼10%) diminished lung function compared to that predicted by age, sex, and race, but other activities were less consistent in direction and magnitude. This relationship has previously been documented, but with hours per day worked as the explanatory variable.[5] Of note, the second tertile of years worked on an IHO shows the greatest decrements in lung function. This may be due to the healthy worker effect, where those who continue working in the industry the longest (third tertile) are the healthiest and can withstand year-after-year exposures to dust and microbes. Others have previously documented improved pulmonary function in IHO workers as they age,[16] and also attributed their findings to healthy worker bias.[38] The heathy worker effect may also explain the unexpected association between pesticide application and improved lung function as measured by FEV_1_/FVC.

While we did not capture data on those quitting IHOs, a prior study found that those who those who left IHO work had lower FEV_1_ measurements than those who remained employed, and the odds of quitting pig operations increased for workers on farms with greater than 400 head of swine.[39] In another population, Donham *et al*. found that baseline

FVC measurements were within 95% of predicted value,[21] an observation that was corroborated in this cohort (**Table 2**), also pointing to healthy worker populations on IHOs.

### Longitudinal analyses were largely null

Unexpectedly, longitudinal analyses did not produce many meaningful results. This is perhaps because of the time scale chosen (once every two weeks), due to the limited measurements of the devices used, or lung function impairment that is too early to detect, as was seen in a study by Schwartz *et al*.[40] It is unlikely that there is no true association between IHO exposures and worker lung function as this has been well documented in prior studies.[41]

We did observe, however, a −0.3 L estimated difference in FEV_1_ during weeks when workers wore any PPE consistently versus those weeks when they did not, which may be explained by (1) improper use leading to a sense of security and thus allowing higher exposures, (2) re-exposure from previously worn, unclean equipment, and/or (3) confounding by work activities associated with PPE usage. This finding led us to conduct a series of *post-hoc* analyses.

### PPE *post-hoc* analyses

Contrary to our hypothesis, in our *post-hoc* analyses we found that coveralls and facemasks were worn less often when workers experienced worse barn conditions and had more pig contact. This may be explained by workers wanting to remove or not don constricting, cumbersome, sweaty, and hot PPE during tasks that may already be uncomfortable. Workers did wear coveralls more consistently as cleaning activities increased, which may be explained by training from an extension office or chemical supplier[36] on the need to protect themselves from chemicals. However, wearing unwashed coveralls may lead to re-exposure of these potentially dangerous substances. Further, at some IHOs, coveralls are required to be worn to protect pigs from humans – especially during the time of data collection which was amid a PEDV outbreak – whereas facemasks and eye protection are not.

Since prior literature has shown face equipment to be protective,[42, 43] training and educational interventions for workers performing increased numbers of dusty or dirty tasks may be a priority for future research. In addition, IHO owners may consider ways to improve workplace conditions (*e*.*g*., better ventilation, dust control measures, fewer animals, hoop barns) so that they are conducive to PPE usage. Given COVID-19, the H1N1 “swine flu” pandemic, our knowledge of antimicrobial resistant pathogens, and increasing awareness about how food systems are linked to the spread of emerging infectious diseases, PPE adherence is particularly important to limit the transmission of diseases off operations[44-47] to protect not only worker but community health.

### Design considerations

This cohort of 103 IHO workers is a non-random, self-selected group, which may lead to potential selection bias arising from differences in those who participated versus those who did not (*e*.*g*., health status, understanding of exposure issues, interest in protecting themselves from work hazards). However, the population is believed to represent the occupational population demographics from the area from which they were enrolled including low-income, minorities, and those living with elder relatives. Further limitations in generalizability include the possible differences between warmer climates (North Carolina) and colder climates, where ventilation systems may differ. Our study was also limited by healthy worker effect bias as detailed above.

### Strengths

While we were unable to use the PEFr measurements, the use of the Piko-1 spirometer to capture FEV_1_ readings was a strength. This easy-to-use handheld device allowed more measurements from workers than had a gold standard Koko tabletop device been used (**Tables 1** and **2**). The Piko-1 was also able to capture more smokers than the Koko. Of the smokers, 12 of 13 had Piko-1 measurements at baseline, while only 5 of 13 had Koko (**Table 1**). Considering that the Piko-1 is not a NIOSH gold standard instrument, it is worth noting that when ATS/NIOSH acceptability criteria are applied to these measurements, the resulting point estimates and confidence intervals were similar to those without these criteria applied (data not shown).

Another strength of this work is the partnership with REACH. This study would not have been possible without community-based participatory research and the community-driven questions they provided. The organization was key to enrolling participants and maintaining an ongoing relationship with IHO workers. The enrollment and high retention rate of hard-to-reach non-white male and female workers performing the day-to-day operations on-IHO is rare.

## Conclusions

In this study we found that healthy IHO workers in North Carolina, the longer a person worked on any IHO the worse their lung function became, as measured by spirometry. It also showed that quality FEV_1_ data can be collected through the use of a handheld, portable asthma-tracking device that does not require a NIOSH-trained technician to be on-site to operate. Further, PPE usage was associated with declines in the respiratory health of IHO workers but is believed to be strongly associated with work activities, where people use face protection less consistently as they perform tasks that involve increased contact with pigs and worsening barn conditions, but wear coveralls more consistently when applying chemicals at work. This may lead to re-exposure from chemicals on coveralls. More research is needed to determine what kinds of masks and coveralls are being used, how they are cleaned, what guidance is being provided to workers, and if interventions in barn conditions could increase compliance and help protect lung function.

## Data Availability

Due to the exceedingly sensitive nature of this data, it will not be made publicly available. For questions regarding access, please contact Dr. Christopher Heaney.

## ACKNOWLEDGEMENTS

This study would not have been possible without a strong partnership between researchers and North Carolinian community-based organization members who have fostered the trust of too often marginalized and at-risk community members. The authors deeply thank the workers who participated in this study.

This manuscript is dedicated to the memory of Dr. Steve Wing, who helped conceive the design and analytical framework for this cohort study.

