## Supplemental Material for "The use of personal protective equipment during common industrial hog operation work activities and acute lung function changes in a prospective worker cohort, North Carolina, USA"

**Table S1.** Classification of adverse respiratory outcomes based on spirometry measurements at baseline within an industrial hog operation worker cohort, North Carolina, 2013-2014.

| Characteristic | *n* (%) |
| --- | --- |
| Global Initiative for Chronic Obstructive Lung Disease (GOLD)[35] (via Koko) |  |
| Obstructive |  |
| No | 68 (99) |
| Yes | 1 (1) |
| Restrictive |  |
| No | 68 (99) |
| Yes | 1 (1) |
| Lower limit of normal (LLN) |  |
| Obstructive (via Koko) |  |
| No | 67 (100) |
| Yes | 0 (0) |
| Obstructive (via Piko-1) |  |
| No | 82 (89) |
| Yes | 10 (11) |
| Restrictive (via Koko) |  |
| No | 67 (100) |
| Yes | 0 (0) |

Note. FEV_1_ = forced expiratory volume in one second; PEFr = peak expiratory flow rate. Normal = FEV_1_ and FVC above 80% predicted (LLN); FEV_1_/FVC ratio above 0.70 (GOLD). Obstructive = FEV_1_ below 80% predicted (LLN); FEV_1_/FVC ratio below 0.70 (GOLD). Restrictive = FVC below 80% predicted (LLN); FEV_1_/FVC ratio above 0.70 (GOLD).

**Table S2.** Crude relationship between occupational activities and spirometry measurements at baseline within an industrial hog operation worker cohort, North Carolina, 2013-2014 using GLM clustered at the household level.

|  | % Predicted FEV_1_*^a^* | | | % Predicted FVC*^a^* | % Predicted FEV_1_/FVC*^a^* | % Predicted FEV_1_*^b^* | |
| --- | --- | --- | --- | --- | --- | --- | --- |
| Characteristic | *n* | β (95% CI) | β (95% CI) | | β (95% CI) | *n* | β (95% CI) |
| Eye, nose, or throat symptoms | 64 | 4.5 (-6.1, 15.1) | 3.6 (-6.5, 13.6) | | -0.4 (-3.3, 2.4) | 89 | 5.4 (-6.2, 17.1) |
| Any allergies | 66 | -11.9 (-26.2, 2.4) | -12.1 (-25.9, 1.7) | | 0.3 (-4.4, 5.0) | 91 | -1.5 (-23.6, 20.5) |
| Doctor-diagnosed asthma | 67 | -10.9 (-23.3, 1.5) | -9.4 (-23.0, 4.2) | | -1.6 (-5.7, 2.6) | 92 | -10.9 (-20.7, -1.1) |

Note. FEV_1_ = forced expiratory volume in one second; FVC = forced vital capacity; CI = confidence interval.

*a.* Performed on a Koko spirometer.

*b.* Performed on a Piko-1 spirometer.

**Table S3.** Hour of test (continuous), current cigarette smoking (binary), and interviewer (dummy) adjusted relationship between occupational activities and spirometry measurements at baseline within an industrial hog operation worker cohort, North Carolina, 2013-2014 using GLM clustered at the household level.

|  |  | % Predicted FEV_1_*^a^* | | % Predicted FVC*^a^* | % Predicted FEV_1_/FVC*^a^* | % Predicted FEV_1_*^b^* | |
| --- | --- | --- | --- | --- | --- | --- | --- |
| Characteristic | *n* | β (95% CI) | β (95% CI) | | β (95% CI) | *n* | β (95% CI) |
| Eye, nose, or throat symptoms | 62 | 6.9 (-3.3, 17.0) | 5.6 (-3.7, 14.8) | | 0.1 (-3.1, 3.2) | 69 | 3.3 (-5.7, 12.3) |
| Any allergies | 63 | -14.7 (-26.6, -2.9) | -14.3 (-26.1, -2.5) | | 0.1 (-4.0, 4.2) | 70 | -6.8 (-25.7, 12.2) |
| Doctor-diagnosed asthma | 63 | -9.7 (-28.1, 8.7) | -8.3 (-27.1, 10.5) | | -1.6 (-4.7, 1.5) | 70 | -13.3 (-30.0, 3.3) |

Note. FEV_1_ = forced expiratory volume in one second; FVC = forced vital capacity; CI = confidence interval.

*a.* Performed on a Koko spirometer.

*b.* Performed on a Piko-1 spirometer.

**Table S4.** Adjusted baseline relationship between reported work exposures and measured lung function within an industrial hog operation worker cohort, North Carolina, 2013-2014.

|  |  | % Predicted FEV_1_ | | % Predicted FVC | | % Predicted FEV_1_/FVC*^a^* | | % Predicted FEV_1_*^b^* | | |
| --- | --- | --- | --- | --- | --- | --- | --- | --- | --- | --- |
| Characteristic | *n* | β (95% CI) | *p*-value | β (95% CI) | *p*-value | β (95% CI) | *p*-value | *n* | β (95% CI) | *p*-value |
| Have you ever |  |  |  |  |  |  |  |  |  |  |
| Given pigs shots and/or antibiotics | 63 | 1.0 (-8.6, 10.7) |  | -0.8 (-9.6, 8.1) |  | 2.8 (-0.5, 6.0) |  | 70 | 3.8 (-8.3, 15.9) |  |
| Drawn pig blood | 63 | 5.5 (-16.9, 27.9) |  | 1.9 (-15.3, 18.9) |  | 1.6 (-3.5, 6.8) |  | 70 | -0.4 (-14.8, 14.0) |  |
| Handled pig manure | 63 | 3.6 (-5.6, 12.7) |  | 4.0 (-6.0, 13.9) |  | -0.9 (-4.3, 2.5) |  | 69 | -2.9 (-13.7, 7.8) |  |
| Applied pesticides in or around the barns | 63 | 0.5 (-7.1, 8.2) |  | -4.2 (-12.7, 4.3) |  | 5.2 (1.7, 8.6) |  | 70 | -1.1 (-9.4, 7.2) |  |
| Washed work clothes with household laundry | 62 | 4.1 (-7.7, 15.9) |  | 2.5 (-9.2, 14.2) |  | 0.8 (-3.8, 5.3) |  | 69 | -1.1 (-15.0, 12.7) |  |
| Do you typically |  |  |  |  |  |  |  |  |  |  |
| Work exclusively in sow, nursery, and/or farrow barns | 61 | -0.2 (-9.1, 8.8) |  | -0.7 (-9.3, 7.9) |  | 1.0 (-2.2, 4.2) |  | 68 | -5.8 (-15.0, 3.5) |  |
| Work exclusively in feeder and/or finisher barns | 61 | -3.2 (-12.5, 6.0) |  | -1.4 (-10.8, 8.0) |  | -1.7 (-6.3, 3.0) |  | 68 | 1.8 (-6.9, 10.6) |  |
| Always wear a mask and bodysuit and eye protection | 62 | 4.1 (-6.5, 14.6) |  | 4.1 (-8.6, 16.7) |  | -0.4 (-4.5, 3.7) |  | 69 | 5.1 (-3.9, 14.1) |  |
| Work 7 days per week | 63 | 0.01 (-8.7, 8.7) |  | 0.4 (-8.9, 9.8) |  | -0.4 (-4.1, 3.2) |  | 70 | 1.3 (-7.0, 9.6) |  |
| Spend 100% of time at work in direct contact with hogs | 63 | -6.2 (-14.9, 2.4) |  | -6.8 (-15.2, 1.7) |  | 0.9 (-2.6, 4.4) |  | 70 | -5.9 (-15.6, 3.8) |  |
| Years worked on any IHO | 59 |  |  |  |  |  |  | 64 |  |  |
| Tertile 1 (1-5 years) |  | Ref (0.0) |  | Ref (0.0) |  | Ref (0.0) |  |  | Ref (0.0) |  |
| Tertile 2 (6-10 years) |  | -7.0 (-16.7, 2.7) |  | -5.7 (-15.9, 4.6) |  | -1.6 (-7.3, 4.0) |  |  | 5.4 (-7.1, 17.9) |  |
| Tertile 3 (11-27 years) |  | -9.0 (-19.4, 1.4) |  | -10.5 (-21.3, 0.4) |  | 0.7 (-2.6, 4.0) |  |  | -5.9 (-17.2, 5.5) |  |
| Trend |  |  | 0.1 |  | 0.1 |  | 0.7 |  |  | 0.4 |
| Percent of life working on any IHO | 59 |  |  |  |  |  |  | 64 |  |  |
| Tertile 1 (2.4-11.6%) |  | Ref (0.0) |  | Ref (0.0) |  | Ref (0.0) |  |  | Ref (0.0) |  |
| Tertile 2 (11.7-26.3%) |  | -9.8 (-20.8, 1.2) |  | -4.3 (-16.5, 8.0) |  | -6.1 (-11.6, -0.7) |  |  | -7.0 (-19.8, 5.7) |  |
| Tertile 3 (26.4-51.9%) |  | -4.2 (-14.3, 6.0) |  | -3.9 (-15.3, 7.5) |  | -1.5 (-5.4, 2.3) |  |  | -7.2 (-18.3, 4.0) |  |
| Trend |  |  | 0.6 |  | 0.5 |  | 0.7 |  |  | 0.2 |

Note. FEV_1_ = forced expiratory volume in one second; FVC = forced vital capacity; CI = confidence interval. IHO = industrial hog operation. All estimates made using GLM clustered at the household level and were adjusted for hour of test (continuous), current smoker (binary), and interviewer (dummy).

*a.* Performed on a Koko spirometer.

*b.* Performed on a Piko-1 spirometer.

**Table S5.** Relationship between reported exposure scores and crude and adjusted FEV_1_ measurements over time within an industrial hog operation worker cohort, North Carolina, 2013-2014 using fixed-effects regression.

|  | Crude FEV_1_ (L) | | | Adjusted*^a^* FEV_1_ (L) | | |
| --- | --- | --- | --- | --- | --- | --- |
| Characteristic in the past week | visits (workers)*^b^* | β (95% CI) | *p*-value | visits (workers)*^b^* | β (95% CI) | *p*-value |
| Barn conditions score*^c^* | 693 (99) |  |  | 684 (99) |  |  |
| 0 |  | Ref (0.0) |  |  | Ref (0.0) |  |
| 1 |  | -0.02 (-0.2, 0.1) |  |  | 0.02 (-0.1, 0.2) |  |
| 2 |  | -0.2 (-0.4, 0.002) |  |  | -0.2 (-0.4, 0.04) |  |
| 3 or 4 |  | 0.1 (-0.3, 0.4) |  |  | 0.03 (-0.3, 0.4) |  |
| Trend |  |  | 0.3 |  |  | 0.4 |
| Cleaning score*^d^* | 717 (99) |  |  | 708 (99) |  |  |
| 0 |  | Ref (0.0) |  |  | Ref (0.0) |  |
| 1 |  | 0.01 (-0.2, 0.2) |  |  | 0.04 (-0.1, 0.2) |  |
| 2 |  | 0.1 (-0.1, 0.2) |  |  | 0.1 (-0.1, 0.2) |  |
| 3 or 4 |  | 0.2 (-0.03, 0.4) |  |  | 0.1 (-0.1, 0.3) |  |
| Trend |  |  | 0.1 |  |  | 0.3 |
| Pig contact score*^e^* | 715 (100) |  |  | 705 (100) |  |  |
| 0 |  | Ref (0.0) |  |  | Ref (0.0) |  |
| 1 |  | -0.1 (-0.2, 0.1) |  |  | -0.1 (-0.3, 0.1) |  |
| 2 |  | -0.1 (-0.3, 0.1) |  |  | -0.1 (-0.3, 0.1) |  |
| Trend |  |  | 0.2 |  |  | 0.2 |
| Score components*^f^* | 680 (99) |  |  | 672 (99) |  |  |
| 0 or 1 |  | Ref (0.0) |  |  | Ref (0.0) |  |
| 2 or 3 |  | -0.1 (-0.2, 0.1) |  |  | -0.1 (-0.2, 0.1) |  |
| 4, 5, or 6 |  | 0.01 (-0.2, 0.2) |  |  | -0.01 (-0.2, 0.2) |  |
| Trend |  |  | 0.8 |  |  | 0.9 |
| PPE score*^g^* | 712 (100) |  |  | 703 (100) |  |  |
| 0 |  | Ref (0.0) |  |  | Ref (0.0) |  |
| 1 |  | -0.3 (-0.6, -0.1) |  |  | -0.3 (-0.6, -0.03) |  |
| 2 |  | -0.4 (-0.7, -0.1) |  |  | -0.4 (-0.8, -0.1) |  |
| 3 |  | -0.3 (-0.7, 0.04) |  |  | -0.4 (-0.8, -0.1) |  |
| Trend |  |  | 0.3 |  |  | 0.05 |
| Number of times washed hands per shift | 717 (100) |  |  | 707 (100) |  |  |
| Tertile 1 (0-6) |  | Ref (0.0) |  |  | Ref (0.0) |  |
| Tertile 2 (7-10) |  | 0.04 (-0.1, 0.2) |  |  | -0.03 (-0.2, 0.1) |  |
| Tertile 3 (11-50) |  | 0.003 (-0.2, 0.2) |  |  | 0.04 (-0.2, 0.3) |  |
| Trend |  |  | 0.8 |  |  | 0.9 |

Note. FEV_1_ = forced expiratory volume in one second; CI = confidence interval; PPE = personal protection equipment.

*a.* Hour of test (dummy), month of test (dummy), smoked in the past 12 hours (binary), and interviewer (dummy)-adjusted relationship between reported exposure scores and spirometry measurements using fixed-effects regression.

*b.* The number of observations equals the number of individual visits (1-8) for the number of persons (i.e., groups) with both a response to the symptom question and a Piko-1 spirometry test result.

*c.* Sum of extreme temperature (yes=1, no=0), extreme malodor (yes=1, no=0), extreme dust (yes=1, no=0), vents off (yes=1, no=0), and a new herd entering the barn(s) (yes=1, no=0)

*d.* Sum of used cleaning chemicals (yes=1, no=0), used pesticides (yes=1, no=0), pressure washed (yes=1, no=0), and used a torch (yes=1, no=0)

*e.* Sum of gave pigs shots (yes=1, no=0) and gave pigs medicine (yes=1, no=0)

*f.* Sum of all above components, with a possible total of 0 to 11.

*g.* Sum of consistently (≥80% of the time at work) wore the following: mask (yes=1, no=0), glasses (yes=1, no=0), and bodysuit/coveralls (yes=1, no=0).

**Table S6.** Crude longitudinal relationship between reported personal protective equipment (PPE) and measured lung function within an industrial hog operation (IHO) worker cohort, North Carolina, 2013-2014, using fixed-effects regression.

| In the past week | FEV_1_ (L) | | | PEFr (L/s) | |
| --- | --- | --- | --- | --- | --- |
|  | visits (workers)*^a^* | ß (95% CI) | visits (workers)*^a^* | | ß (95% CI) |
| Used body protection consistently*^b^* | 719 (101) | -0.2 (-0.4, -0.03) | 719 (101) | | -0.5 (-1.1, 0.01) |
| Used face protection*^c^* consistently | 713 (100) | -0.1 (-0.2, 0.1) | 713 (100) | | -0.01 (-0.5, 0.4) |

*a.* The number of observations equals the number of individual visits (1-8) for the number of persons (i.e., groups) with both a response to the personal protective equipment question and a Piko-1 spirometry test result.

*b.* Consistently defined as ≥80% of the time at work.

*c.* Face protection = either reported mask or eye protection.

**Table S7.** Cross tabulations of personal protective equipment use and reported symptoms within an industrial hog operation worker cohort, North Carolina, 2013-2014.

|  | Body protection | | | Face protection | | |
| --- | --- | --- | --- | --- | --- | --- |
| Reported in the past week | <80% | ≥80% | *p*-value (Pearson *X*^2^) | <80% | ≥80% | *p*-value (Pearson *X*^2^) |
| At least one respiratory symptom*^a^*, *n (%)* |  |  |  |  |  |  |
| No | 148 (95) | 541 (94) | 0.7 | 341 (95) | 342 (94) | 0.6 |
| Yes | 8 (5) | 34 (6) |  | 19 (5) | 23 (6) |  |
| At least one symptom interfered with sleep*^b^*, *n (%)* |  |  |  |  |  |  |
| No | 146 (97) | 544 (96) | 0.5 | 344 (97) | 340 (95) | 0.1 |
| Yes | 4 (3) | 22 (4) |  | 9 (3) | 17 (5) |  |
| Sneezing, *n (%)* |  |  |  |  |  |  |
| No | 152 (97) | 564 (98) | 0.9 | 357 (99) | 353 (96) | 0.1 |
| Yes | 4 (3) | 14 (2) |  | 5 (1) | 13 (4) |  |
| Headache, *n (%)* |  |  |  |  |  |  |
| No | 151 (97) | 568 (98) | 0.3 | 356 (98) | 357 (98) | 0.5 |
| Yes | 5 (3) | 10 (1) |  | 6 (2) | 9 (2) |  |
| Eye or nose symptoms, *n (%)* |  |  |  |  |  |  |
| No | 153 (98) | 567 (98) | 1.0 | 356 (98) | 358 (98) | 0.6 |
| Yes | 3 (2) | 11 (2) |  | 6 (2) | 8 (2) |  |

Note.

*a.* Excessive coughing, runny nose, difficulty breathing, or sore throat.

*b.* Any sleep symptoms reported, waking from sleep due to coughing, waking from sleep due to wheezing, or waking from sleep due to phlegm.
